# COSTS ASSOCIATED WITH TRANSRADIAL ACCESS AND SAME-DAY DISCHARGE AFTER PERCUTANEOUS CORONARY INTERVENTION: A SYSTEMATIC REVIEW AND META-ANALYSIS

**DOI:** 10.1101/2020.08.18.20177063

**Authors:** Hemant Kulkarni, Manoj Thangam, Samuel Lindner, Christian McNeely, Amit P. Amin

## Abstract

**Introduction:** Transradial access for PCI (TRI) along with same day discharge (SDD) is associated with varying estimates of cost savings depending on the population studied, the clinical scenario and application to low-risk vs high-risk patients. A summary estimate of the true cost savings of TRI and SDD are unknown.

**Methods:** We searched the PubMed, EMBASE®, CINAHL® and Google Scholar® databases for published studies on hospitalization costs of TRI and SDD. Primary outcome of interest in all included studies was the cost saving with TRI (or SDD), inflation-corrected US$ 2018 values using the medical consumer price index. For meta-analytic synthesis, we used Hedges’ summary estimate (g) in a random-effects framework of the DerSimonian and Laird model, with inverse variance weights. Heterogeneity was quantified using the 12 statistic.

**Results:** The cost savings of TRI from four US studies reported a consistent and significant cost saving associated with TRI after accounting for currency inflation, of US$ 992 (95% CI US$ 8501,134). The cost savings of SDD from six US studies, after inflation-correcting to the year 2018, were US$ 3,567.58 (95% CI US$ 2,303 –4,832).

**Conclusions:** In conclusion, this meta-analysis demonstrates that TRI and SDD are associated with mean cost reductions of by approximately US$1,000/patient and US$ 3,600/patient, respectively, albeit with wide heterogeneity in the cost estimates. When combined with the safety of TRI and SDD, this meta-analysis underscores the value of combining TRI and SDD pathways and calls for a wide-ranging practice change in the direction of TRI and SDD.

## INTRODUCTION

The practice of percutaneous coronary intervention (PCI) in the United States is evolving. Over 25 years ago, Campeau in Canada contemporaneous with Kiemeneij and Laarman in The Netherlands, began explorations into the possibility of using transradial access.^1^–^7^ Since then rapid advances in this technique and related devices have catapulted transradial intervention (TRI) to an internationally accepted benchmark and a clinically viable approach to PCIs.^8^–^11^ A series of studies have demonstrated the superiority of TRI compared to conventional transfemoral intervention (TFI) in terms of reduced risk of adverse outcomes and shorter lengths of hospital stays.^12^–^23^ A recent Scientific Statement from the American Heart Association highlights these advantages and recommends TRI even in patients with acute coronary syndrome (ACS).^24^ Additionally, over a decade ago, Society for Cardiovascular Angiography and Interventions (SCAI) published an expert consensus in the form a decision matrix to facilitate same-day discharge (SDD) after PCI.^25^ Being especially suited for TRI, this SDD approach is highly attractive to patients, physicians and hospitals.^26^–^28^ A recent metaanalysis has shown that in patients with stable coronary artery disease, SDD is not associated with adverse outcomes. The SDD approach has thus led to reduced practice variations around post-PCI care without affecting the rates of adverse outcomes after PCI.^28^–^31^

Both TRI and SDD have added economic benefits and have been shown to reduce costs associated with PCI.^1,32^”^40^ From the perspective of the hospitals, better resource utilization and shorter duration of hospital stay can yield substantial reductions in hospitalization costs. While there is a paucity of controlled trials that specifically address hospitalization costs in the context of TRI and SDD, several observational studies have demostrated reduced hospitalization costs in this regard.^15, 32–34, 36, 41–43^ However, these studies have provided varying estimates of cost savings depending on the population studied, the clinical scenario and application to low-risk vs high-risk patients. Accordingly, the true cost savings of TRI and SDD have not been defined. To address this knowledge gap, we report our results from a systematic review and meta-analytic synthesis of the published literature on the estimated economic impact of TRI and SDD in the United States. Our objectives were twofold: to determine a meta-analytic summary estimate of the cost savings associated with TRI and SDD; and to investigate the heterogeneity across studies in cost-savings associated with TRI or SDD.

## METHODS

### Data extraction

We searched the PubMed, EMBASE®, CINAHL® and Google Scholar® databases for published studies on hospitalization costs of TRI and SDD. Figure 1 shows the flowchart used to include studies into this systematic review and meta-analysis. A combination of literature search and snowballing methods were used to include potential studies into this review. Each included study was independently reviewed by the two coauthors (AA and HK) for completeness and to extract the information needed for meta-analysis. We included all published studies related to cost savings of TRI or SDD in the systematic review but for the meta-analyses we restricted to the US studies only to avoid comparisons across different healthcare models

**Figure 1.**
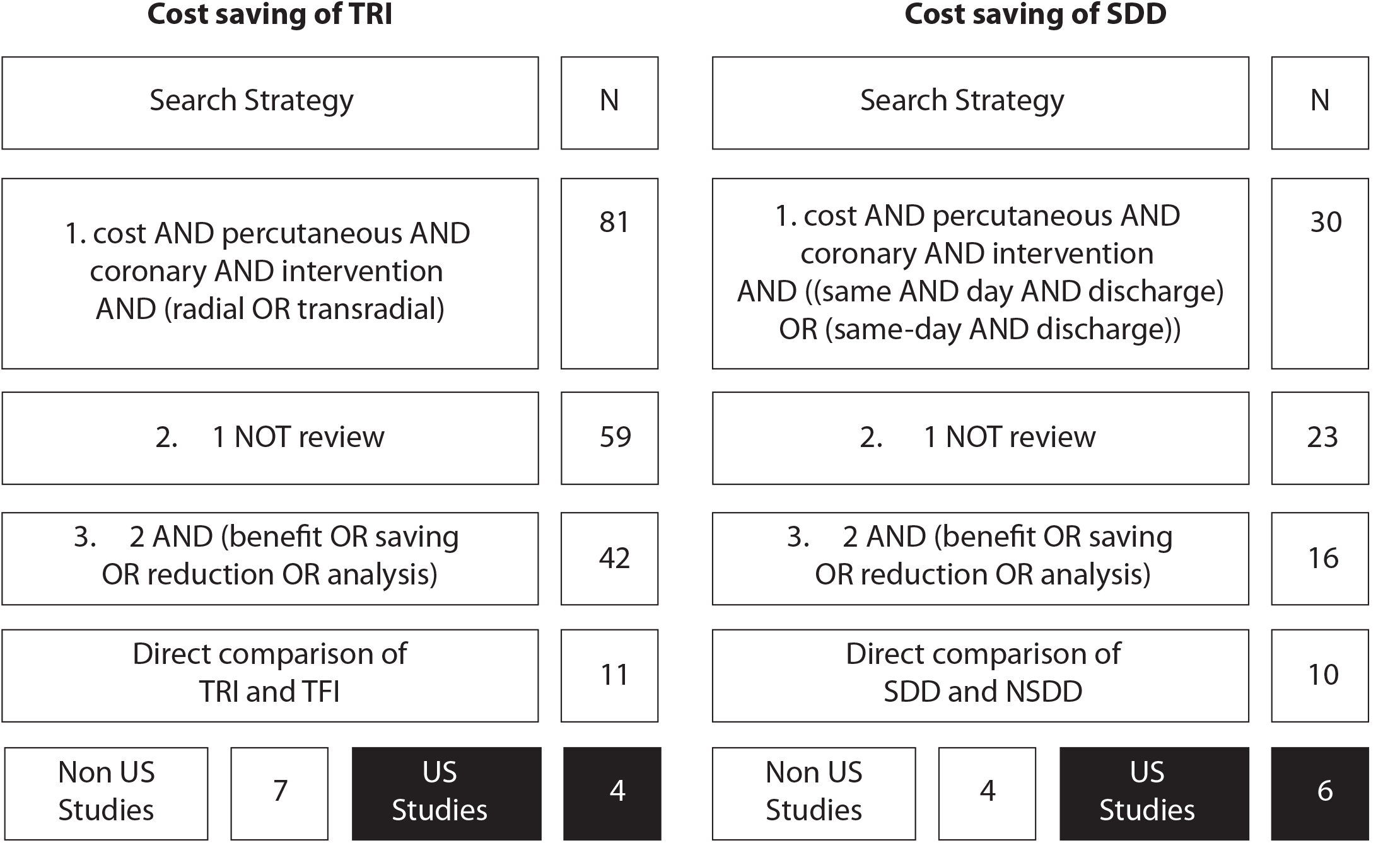
Protocol for including studies in this systematic review and meta-analysis. Black boxes indicate studies that were included in the meta-analyses. N, number of articles.

### Statistical analysis

Primary outcome of interest in all included studies was the absolute cost savings associated with TRI or SDD. Since the studies have been published at different times, we first converted the cost estimates reported by individual studies into inflation-corrected 2018 United States dollar (US$) values using the medical consumer price index.^44,45^ If a study reported the inflation-corrected year for cost estimates, then the 2018 US$ value was obtained by appropriate correction to the reported year. Otherwise, midpoint of the data collection period was used as the proxy year for the cost estimates and the inflation correction was applied to this proxy year.

For meta-analytic synthesis, we used Hedges’ summary estimate (g)^46^ in a random effects framework of the DerSimonian and Laird model^47^. Inverse variance weights were used to compute the summary estimates. Heterogeneity was quantified using the I^2^ statistic.^48^ Sources of heterogeneity were investigated using two approaches: first, we constructed the Baujat plots^49^ to investigate the simultaneous influence of each study on summary estimate and heterogeneity. Second, we used meta-regression in an univariable fashion to estimate the I^2^ explained by putative confounders. Statistical analyses were conducted using the metan software package^50^ under STATA 12.0 (College Station, Texas). Statistical significance was tested at a type 1 error rate of 0.05.

## RESULTS

### Costs of TRI versus TFI in the United States

#### Initial economic studies related to TRI versus TFI

In 1996, Mann et al published the first cost-effectiveness study of TRI. The primary outcome in this study was the hospital charge of TRI as compared to TFI and it was found that in a randomized, controlled setting of 152 PCIs, the hospital charge for TRI procedures was significantly lower than that for TFI ($14,374 versus $15,796).^51^ The same group pf investigators published another randomized trial of ACS patients in 1998 and reported that the TRI approach was associated with lower hospital charges as compared to TFI ($20,476 versus $23,389) in ACS patients also.^52^ For over a decade after these initial reports, economic evaluation of TRI was not undertaken in the United States. Much of the focus during this time was on demonstrating that the complications and outcomes of TRI versus TFI are either comparable or improved.

#### Recent economic studies of TRI versus TFI

A prior meta-analysis^24^ published in 2012 on the heels of an earlier meta-analytic cost-benefit analysis^53^ has detailed the economic benefits of TRI over TFI. For example, it has been shown that the lower complication rates and shorter hemostasis time associated with radial access are sufficient to offset the comparable (or potentially longer) procedural time and higher crossover rates such that there is a net estimated direct cost saving of $275 per patient.^53^ This estimate was reflective of the costs saved by way of reduced complications which are costly in the context of PCI.^53^

A year after this meta-analysis was published, Safley et al^54^ used a propensity-matched analysis from a large, nationally representative repository and estimated the per-patient saving of $533 attributable to TRI. The cost savings associated with TRI in this study, primarily stemmed from a reduced post-PCI length of stay among TRI patients. Notably, the cost savings were directly proportional to the estimated bleeding risk such that in patients with high predicted probability of bleeding there could be a $1046 cost saving with TRI. Since this study, this group of investigators has published two additional studies based on patients in the NCDR registry (5 hospitals, n = 7,121) and Medicare beneficiaries (n = 279,987) which have again showed statistically significant cost savings associated with TRI.^36,55^ One of these two studies^36^, also demonstrated a corroborating finding that cost savings attributable to TRI can be greater in patients with a high predicted risk of bleeding ($1621).

#### Meta-analysis of US studies on the cost savings of TRI

Figure 2A shows a forest plot of the four US studies on TRI-related cost savings that were included in meta-analyses. Table 1 shows that these studies came from widely varying patient sources – single center, Premier Healthcare database, NCDR CathPCI for 5 hospitals and Medicare beneficiaries. These studies generally reported a consistent and significant cost saving associated with TRI that ranged from US$553 – US$916 per PCI. After accounting for currency inflation, the results from random-effects meta-analysis showed that the estimated, average, adjusted cost savings attributable to TRI in 2018 were US$ 992.30 (95% CI 850.36 – 1134.23). Also, despite varying sources of patient information and largely varying sample sizes, these studies were still statistically homogeneous (I^2^ = 0, p 0.674) indicating that the summary estimate of cost saving obtained from the meta-analysis was reliable. This homogeneity is evident also from the Baujat plot (Figure 2B). As shown in the Baujat plot, the overall Q statistic of 1.54 was largely contributed by one study^54^ that did not impact the summary estimate of cost saving while the remaining three studies did not contribute to heterogeneity substantially.

**Figure 2.**
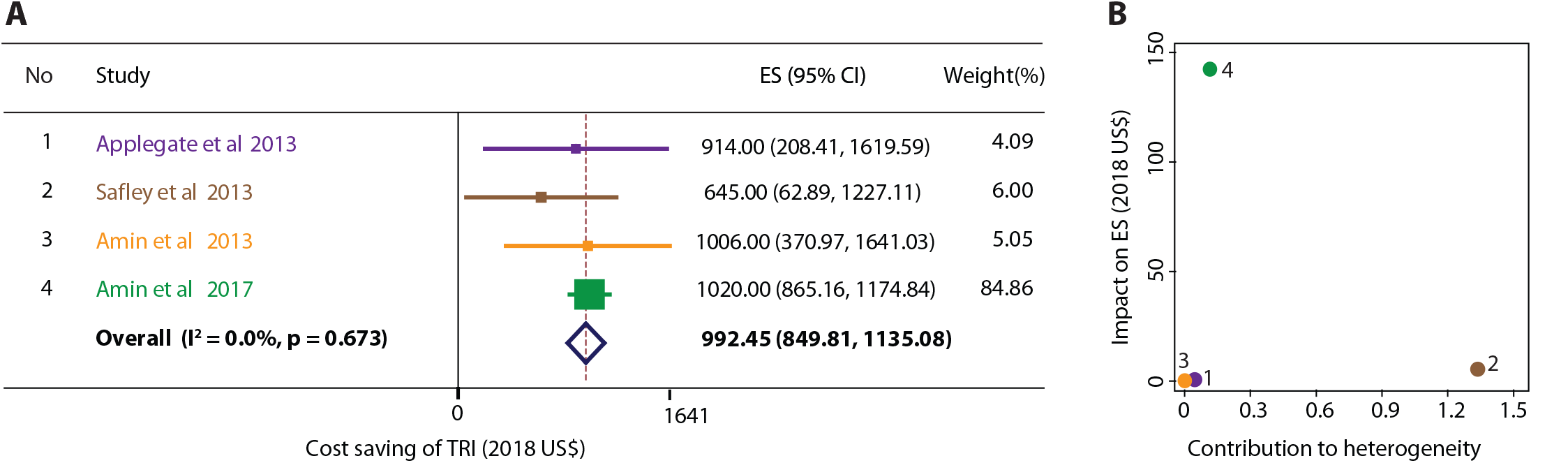
Meta-analysis of the cost savings of TRI. **(A)** Forest plot of the published studies. Each study is color-coded. Boxes are proportional to the inverse-variance weights, error bars indicate 95% confidence intervals. Diamond represents the summary effect size. (B) Boujat plot for the studies included in meta-analysis. Studies are color-coded and numbered to match the scheme in panel A.

**Table 1.** Characteristics of the studies included in the meta-analysis.

| Study | Year | Type of study | N | Patient Source | Prevalence of TRI | Cost period | Cost of TRI/SDD |  | Cost of TFI/NSDD |  | Inclusion/Exclusion | Study period | Dollar year | Cost Reduction |  | 2018 cost reduction |  |
| --- | --- | --- | --- | --- | --- | --- | --- | --- | --- | --- | --- | --- | --- | --- | --- | --- | --- |
|  |  |  |  |  |  |  | Mean | SD | Mean | SD |  |  |  | Mean | SD | Mean | SD |
| Studies on cost saving of TRI |  |  |  |  |  |  |  |  |  |  |  |  |  |  |  |  |  |
| Applegate et al | 2013 | Observational | 1,140 | Wake Forest Baptist Medical Center, Winston-Salem, NC | 0.5 | Hospitalization | 15,186 | 4,803 | 15,918 | 5,234 | PCIs after Sep 2009, elective only, no STEMI, no CABG, propensity matching | Jan 2009-Dec 2011 | 2010 | 732 | 288 | 914 | 360 |
| Safley et al | 2013 | Observational | 61,509 | Premier Healthcare Database | 0.0909 | Hospitalization | 11,736 | 268 | 12,288 | 393 | All patients, single PCI, single stent, no CS, no data on MI status | 2004-2009 | 2012 | 552 | 254 | 645 | 297 |
| Amin et al | 2013 | Observational | 7,121 | NCDR (5 hospitals) | 0.1712 | Hospitalization | 14,954 | 204 | 15,784 | 161 | PCIs at study centers, single PCI, no CS, no CTO | Jan 2010-Mar 2011 | 2011 | 830 | 267 | 1,006 | 324 |
| Amin et al | 2017 | Observational | 279,987 | Medicare beneficiaries | 0.0904 | Hospitalization | 15,786 | 72 | 16,701 | 41 | All matching claims, no ACS, no CTO, no CS, no CA | Jul 2009-Dec 2012 | 2014 | 915 | 71 | 1,020 | 79 |
| Studies on cost saving of SDD |  |  |  |  |  |  |  |  |  |  |  |  |  |  |  |  |  |
| Amin et al | 2017 | Observational | 279,987 | Medicare beneficiaries | 0.0529 | Hospitalization | 13,256 | 83 | 16,753 | 40 | All matching claims, no ACS, no CTO, no CS, no CA | Jul 2009-Dec 2012 | 2014 | 3,502 | 78 | 3,902 | 86 |
| Amin et al | 2018 | Observational | 672,470 | Premier Healthcare Database | 0.0906 | Hospitalization | Not reported |  |  |  | Elective PCIs | Jan 2006 - Dec 2015 | 2016 | 5,128 | 61 | 5,364 | 64 |
| Amin et al | 2018 | Observational | 1,752 | Single center NCDR CathPCI data | 0.1312 | Hospitalization | Not reported |  |  |  | Elective PCIs | Jul 2009-Sep 2015 | 2016 | 7,331 | 1,481 | 7,668 | 1,549 |
| Rodriguez-Araujo et al | 2018 | Observational | 490 | Single center registry | 0.5 | 30 days | 4,493 | 119 | 7,112 | 267 | Elective, transradial PCIs from RASSADA PCI trial |  | 2018 | 2,619 | 293 | 2,621 | 293 |
| Kwok et al | 2019 | Observational | 324,345 | NRD repository | 0.0157 | 30 days | 15,063 | 109 | 16,968 | 34 | Elective PCIs, no MCS, no CS, no CABG | 2010-2014 | 2012 | 1,905 | 114 | 2,227 | 133 |
| Rymer et al | 2019 | Observational | 2,184 | VA CART repository | 0.33 | 30 days | 23,656 | 574 | 25,878 | 458 | First, unstaged, elective PCI, no weekend adm, no urgent or salvage procedures, no in-lab complications | Oct-2008 - Sep 2016 | 2016 | 1,503 | 383 | 1,572 | 400 |

#### Corroborating evidence from around the world

Table 2 shows the evidence for significant cost savings associated with TRI use as reported by studies around the world. There have been 7 published studies^15,56^”^61^ that have reported cost differential in TRI and TFI – 3 studies from China^58^–^60^ and 1 each from France^56^, Mexico^57^, Poland^15^ and England^61^. Irrespective of the country wherefrom a study was reported, there was a consistent observation of a reduced cost associated with TRI. For international cost savings, the international currency conversion rates, varying payment systems, and different publication years of these studies make it difficult to arrive at a standardized estimate of cost saving that is comparable across the datasets but the consistency of derived estimates of cost savings in these studies is evident. Data presented in these studies again demonstrate reduced rates of complications as the primary driver rather than procedural cost differences. For example, in a small trial, Dziki et al^62^ demonstrated comparable procedural cath lab costs when comparing TRI to TFI. Finally, it is noteworthy that the recent, large study^61^ from England again demonstrated that the TRI costs are even lower in STEMI patients (348£) as compared to stable CAD patients (153£) – a finding that is concordant with the Safley et al^54^ and Amin et al^55^studies from the US implying that the cost savings tend to be higher in high-risk patients. Together these studies corroborate the results of meta-analysis of the US studies presented in Figure 2A.

**Table 2.** Studies on cost savings of TRI from around the world.

| Study | Year | Type of study | N | Country | Prevalence of TRI | Inclusion/Exclusion | Cost Reduction | Currency |
| --- | --- | --- | --- | --- | --- | --- | --- | --- |
| CARAFE | 2001 | RCT | 210 | France | 0.66 | All PCIs, no MI, no CABG, no RHC | 586.5 | FrF |
| Escárcega et al | 2010 | Observational | 174 | Mexico | --- | 131 stable coronary patients, 43 ACS patients | 509.0 | \$ |
| Jin et al | 2016 | Observational | 5,306 | China | 0.885 | Single center, single PCI, no CS, no CTO, no IABP | 8,081.0 | ¥ |
| Koltowski et al | 2016 | RCT | 103 | Poland | 0.5 | STEMI patients from OCEAN RACE trial | 314.0 | € |
| Jin et al | 2017 | Observational | 1,229 | China |  | Elderly patients | 7,495.0 | ¥ |
| Jin et al | 2017 | Observational | 1,392 | China |  | Women only | 7,474.0 | ¥ |
| Mamas et al | 2018 | Observational | 248,228 | England | 0.5 | All indications | 250.6 | £ |
|  |  | Observational | 94,856 | England | 0.5 | Elective/Stable CAD only | 153.9 | £ |
|  |  | Observational | 87,904 | England | 0.5 | NSTEMI/UA only | 282.2 | £ |
|  |  | Observational | 65,468 | England | 0.5 | STEMI only | 348.3 | £ |

### Economic benefits of Same-day discharge after PCI

#### SDD in eligible patients is safe and effective

Abedlaal et al^28^ published a meta-analysis of randomized controlled trials (RCTs) and observational studies that demonstrated a similar clinical outcomes for SDD versus overnight stay. Another meta-analysis^63^ published in 2017 also demonstrated similar findings. In a recently published meta-analysis of 11 studies with 21,687 patients, Lu et al have demonstrated that early and late clinical outcomes in patients discharged on the same day of PCI were comparable to those discharged after an overnight stay. While the clinical value of SDD is thus clearly established^65^, the subsequent reductions in costs of PCI have not been systematically studied and are not clearly established.

#### Meta-analysis of US studies on cost savings of SDD

Six studies ^32, 33, 36, 41, 66, 67^ were eligible for meta-analysis of the cost savings of SDD in the US. Notably, these studies represent research published in the last three years (2017–2019). All the included studies were observational in nature with prevalence of SDD varying widely from 1.6% to 50% (two studies represented a matched comparison of SDD versus NSDD in a 1:1 or 1:2 proportion). Three studies^36,41,66^ reported costs over the duration of hospitalization while the other three studies^32,33,67^ reported cumulative 30-days costs. The reported mean adjusted per-PCI cost savings in these studies ranged from US$ 1,503 to US$ 7,331.

After inflation-correcting to year 2018 and using the random effects meta-analytic framework, we estimated that the mean per-PCI cost savings of SDD were US$ 3,567.58 (95% CI US$ 2,303.39 – US$ 4,831.78, Figure 3A). However, there was a highly significant heterogeneity in the cost savings estimates across the studies (I^2^ = 99.2%, p < 0.001). We therefore investigated the source of this heterogeneity using Baujat plot (Figure 3B). We found that the heterogeneity Q statistic of 617.56 was mainly contributed to by two studies^41,67^ (combined contribution = 79% of overall heterogeneity). Both these studies have large sample sizes and narrow confidence intervals around the estimated cost saving and, as a result, contributed to the overall heterogeneity (Figure 3B). Of these two however, one study^67^ did not have a substantial impact on the summary effect size. When we conducted subgroup meta-analyses based on whether the studies reported hospitalization costs or 30-day cumulative costs, we found that the summary estimate of cost saving was higher for hospitalization costs than that for the 30-day costs (US$ 5,042 versus US$ 2,208). In the subgroup analyses, significant heterogeneity remained for the hospitalization cost studies (I^2^ = 98.9%, p< 0.001) but not for the 30-day cost studies (I^2^ = 55.4%, p 0.106, Figure 3A).

**Figure 3.**
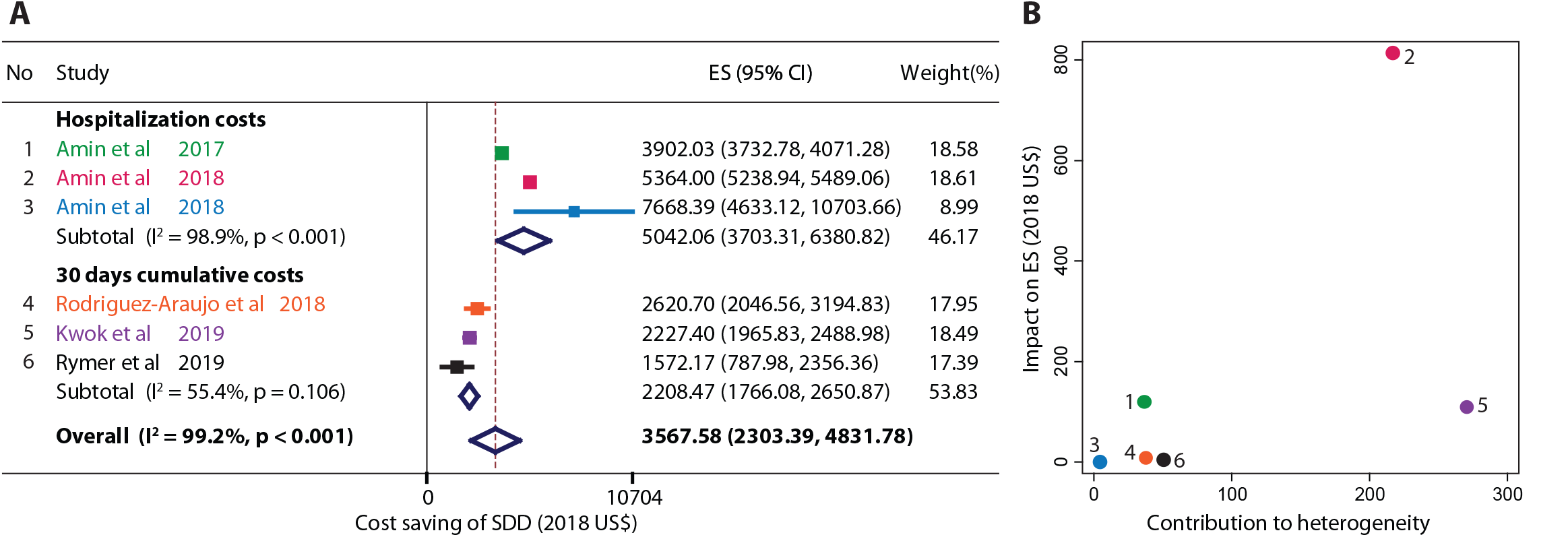
Meta-analysis of the cost savings of SDD. **(A)** Forest plot of the published studies. Each study is color-coded. Boxes are proportional to the inverse-variance weights, error bars indicate 95% confidence intervals. Diamonds indicate the group-specific and overall summary effect size. **(B)** Boujat plot for the studies included in meta-analysis. Studies are color-coded and numbered to match the scheme in panel A.

Using univariable meta-regression we also investigated the potential contribution of cost period, prevalence of SDD, prevalence of TRI, average age, proportion of females and the proportion of elective PCIs on heterogeneity. We found that only cost period (hospitalization costs versus 30-day costs) reached statistical significance (p = 0.029) whereas none of the remaining putative predictors could adequately explain between-study heterogeneity.

#### Combined influence of TRI and SDD on cost savings

Marso and Amin et al^68^ had conjectured that a combination of TRI and SDD could be highly desirable. The Amin et al study^36^ first demonstrated that the cost saving in patients who underwent TRI and SDD as compared to those who underwent TFI and NSDD was US$ 3,689 (95% CI US$ 3,486 – US$3,902) – largest in all pairwise comparisons based on access site and SDD. Next largest cost saving was in patients who underwent TRI and were compared based on whether or not they were discharged same day. The cost saving in TRI-SDD patients was US$ 3,035 (95% CI US$ 2,825 – US$3,273) when compared with TRI-NSDD patients. This comparison is similar to the Rodriguez-Araujo study^33^ which only included transradially accessed elective PCIs and reported a cost reduction of US$ 2,619 (95% CI 2,033 – US$ 3,204). While the small number of studies reporting cost saving of SDD in the context of TRI and the heterogeneity in the reference groups used for estimating the cost savings preclude a formal meta-analysis, these observations point towards substantial cost savings by combining TRI and SDD.

#### Supportive evidence for SDD-based cost savings from around the world

A randomized controlled trial of SDD (the EPOS trial) in TFI patients from The Netherlands (2007) first reported a cost saving of US$ 258 attributable to SDD.^69^ Later, economic analyses of the patients enrolled in the Canadian EASY clinical trial showed that there was a per patient saving of $1,141 when SDD was resorted to.^43^ In 2013, a French study showed a cost saving of €1,074 attributable to ambulatory discharge as compared to conventional PCI.^70^A study from Ontario, Canada found that SDD PCI costs roughly 1,200 Canadian Dollars less than NSDD PCI.^29^ These international studies from various countries together lend further support in favor of a consistent cost saving associated with SDD.

## DISCUSSION

To our knowledge, this is the first systematic review and metaanalyses of the cost savings associated with TRI and SDD in PCI. Our report makes the following key observations: first, TRI was associated with an average cost reduction of US$ 992; second, while the reported estimates of cost savings of SDD have been higher than that of TRI, more homogeneous studies are needed before a reliable estimate of SDD cost savings can be obtained; third, a combination of TRI and SDD is likely to yield significant cost savings for patients undergoing PCI; and lastly, all of these inferences are consistently supported by numerous trials and registries.

According to the Institute of Medicine, an ideal healthcare system should provide care that is safe, effective, efficient, equitable, timely and patient-centered.^71^ In this context, the appropriateness of both TRI and SDD as candidate approaches to PCI care is self-evident. Both approaches are safe (as shown by several previous studies and meta-analyses),^33,72^”^75^ effective as demonstrated by the associated cost savings shown here; efficient (as demonstrated by studies on time demands of PCI options)^43,76^”^78^ and patient-centered (since they both target patient satisfaction)^66^. In this context, it is encouraging to note that the use of TRI in the United States is rapidly increasing. The NCDR CathPCI Registry showed that in 2014 that the prevalence of TRI had risen to 25% of all PCIs while the VA-CART Registry showed a prevalence of 32% of TRI in 2018.^79^,^81^ However, even today most of the patients still undergo TFI. Assuming a total of 900,000 PCIs nationally, even if the TRI rate were to increase modestly from 30% to 50%, then we estimate a national cost saving of 178.5 million US$ annually. As indicated by the studies in this review, this estimate is likely to be even higher (although restricted to elective PCIs only) by increasing use of SDD. Lu et al state^64^ that 57% of the interventionists in the US practice SDD. These practice trends (switching over to the TRI/SDD pathway) need to therefore continue and be bolstered.

The size of both the meta-analyses presented here also points towards a need for more studies in this domain. However, a glaring finding from our analyses is the paucity of randomized, controlled studies on the cost savings of both TRI and SDD (especially the latter). Most of the data that currently exists comes from observational studies. Whereas these observational studies have provided key insights into the econometrics of PCI, measured or unmeasured confounding can remain even if all the studies have rigorously adjusted for as many confounders as possible.^82,83^ In the future, large-scale multicenter randomized studies evaluating the costs of various care pathways (e.g. TRI/SDD) could provide a robust estimate of financial effects of these interventions. Combining TRI and SDD pathways is logical since, as shown by Koutouzis et al^84^, the main driver of SDD is TRI. These studies are essential for wide-ranging practice changes in the direction of TRI and SDD.

Lastly, the reasons for heterogeneity in studies related to SDD cost savings could not be adequately ascertained in this meta-analysis. Perhaps, the duration of cost ascertainment (hospitalization versus 30-day costs) may be a contributor, but it did not fully explain the residual heterogeneity. Other study characteristics such as age distribution, gender representation and TRI prevalence did not significantly contribute to the between-study heterogeneity. More studies and more comprehensive investigations into heterogeneity are needed in future.

## CONCLUSION

There is adequate evidence to indicate that TRI can reduce hospitalization costs of PCI by approximately US$ 1,000 per patient. Published studies indicate that cost savings of SDD are likely to be even higher but substantial heterogeneity exists in estimates of cost savings across studies. More robust data from controlled trials are needed to reliably establish SDD cost savings.

## Data Availability

Not applicable

